# SARS-CoV-2 testing in the community: Testing positive samples with the TaqMan SARS-CoV-2 Mutation Panel to find variants in real-time

**DOI:** 10.1101/2021.11.17.21266297

**Authors:** Fiona Ashford, Angus Best, Steven Dunn, Zahra Ahmed, Henna Siddiqui, Jordan Melville, Samuel Wilkinson, Jeremy Mirza, Nicola Cumley, Joanna Stockton, Jack Ferguson, Lucy Wheatley, Elizabeth Ratcliffe, Anna Casey, Tim Plant, The COVID-19 Genomics UK (COG-UK) Consortium, Joshua Quick, Alex Richter, Nicolas Loman, Alan McNally

## Abstract

Genome sequencing is a powerful tool for identifying SARS-CoV-2 variant lineages, however there can be limitations due to sequence drop-out when used to identify specific key mutations. Recently, Thermo Fisher Scientific have developed genotyping assays to help bridge the gap between testing capacity and sequencing capability to generate real-time genotyping results based on specific variants. Over a 6-week period during the months of April and May 2021, we set out to assess the Thermo Fisher TaqMan Mutation Panel Genotyping Assay, initially for three mutations of concern and then an additional two mutations of concern, against SARS-CoV-2 positive clinical samples and the corresponding COG-UK sequencing data. We demonstrate that genotyping is a powerful in-depth technique for identifying specific mutations, an excellent complement to genome sequencing and has real clinical health value potential allowing laboratories to report and action variants of concern much quicker.

## INTRODUCTION

Viruses mutate and SARS-CoV-2 is no exception. As the COVID-19 pandemic continues around the world, mutations are naturally occurring resulting in the emergence of divergent clusters / variants containing sets of mutations. These clusters/variants have differing prevalence in different geographical regions, likely in response to changing immune profiles of the human population **(1)**. Movement of people enabled by global air travel, allow these variants to spread and mutate further under differing selection pressures. In the United Kingdom, the Alpha variant (B.1.1.7), first identified in Kent, rapidly swept to dominance by December 2020 **(2)**. In April 2021, the Delta variant (B.1.617.2), first identified in India, rapidly outcompeted the Alpha variant to become dominant in a matter of weeks **(3)**. The geographical location where these variants emerged is not relevant **(4)**, rather it is the specific mutations present which greatly impacts virus characteristics including transmissibility and antigenicity, where mutations of significance have been identified in the SARS-CoV-2 Spike Protein of these variants of concern (VOC), that contribute to enhanced transmission and/or immune invasion **(5)**. Other VOC have been identified and characterised, including both Beta (B.1.351 – first identified in South Africa) and Gamma (P.1 – first identified in Brazil) **(5)**.

Previously at the University of Birmingham, we set up a SARS-CoV-2 modular testing facility at the request of the United Kingdom Department of Health and Social Care **(6)**. Our PCR assay of choice was the 3-target design (ORF1ab, S and N genes) TaqPath COVID-19 CE-IVD RT PCR Kit, where target areas are unique to SARS-CoV-2 to reduce detection of other coronaviruses and compensate for virus mutations. Initially, we detected all three genes in SARS-CoV-2 samples, however, during November 2020, we along with other laboratories using the same PCR assay, started to notice a drop-off in the detection of the S-gene and then a rapid rise in this S-gene target failure (SGTF) in early December 2020 **(7)**. From discussions with the University of Birmingham genome sequencing laboratory (as part of the COG-UK Consortium) and as confirmed by other laboratories, it was demonstrated that S-gene did amplify, therefore confirming this gene was still present, but was not being detected using the TaqPath COVID-19 assay. This was attributed to a 6-bp deletion (Δ69/70) in the middle of the S-gene where the fluorescent specific probe binds, thus negating the probe’s ability to fluoresce (**8**). Simultaneously, these findings of a new VOC were reported by our laboratory and multiple testing facilities across the UK, where this finding identified the VOC B.1.1.7 (Alpha variant). Although SGTF identification with the Thermo Fisher TaqPath RT-qPCR assay was not intentional, this observation with this PCR assay allowed us and other COVD-19 testing facilities to use this assay as an accurate epidemiological tool to track the rise, spread and dominance of this VOC in the UK. By April 2021, COVID-19 TaqPath PCR testing facilities, including our laboratory, noted an increasing number of samples without SGTF, where the 3 target genes were amplifying and upon genome sequencing analysis these samples were identified as the Delta (B.1.617.2) variant (**9**).

To better understand viral transmission and evolution and to inform public health responses and vaccine development, genomic sequencing is essential. In March 2020, the COVID-19 Genomics UK Consortium (COG-UK) was created for this purpose (**10**). To date, COG-UK have sequenced over 1,100,000 SARS-CoV-2 samples, providing a vast amount of data to the global COVID-19 response. This yields crucial information about the number of variants circulating in the population and possible lines of transmission, however, sequencing can be timely, costly and in some cases full coverage of the virus is not possible.

Recently, Thermo Fisher have developed genotyping assays to help bridge the gap between testing capacity and sequencing capability to receive results in real-time, so in addition to our modular system for COVID-19 testing, we decided to build into our work-flow the Thermo Fisher TaqMan™ SARS-CoV-2 mutation panel (Figure 1). Here we present our data showing that the TaqMan genotyping assays identify variants in all samples tested with zero failure rate, and that often the assay confirms mutations in a viral isolate that cannot be definitively identified from genome sequence data alone. We conclude that the genotyping assay is an excellent complement to genome sequencing efforts and allows rapid, point-of-testing determination of the presence of any genetic variant for SARS-CoV-2 for which an assay can be designed.

**Figure 1.**
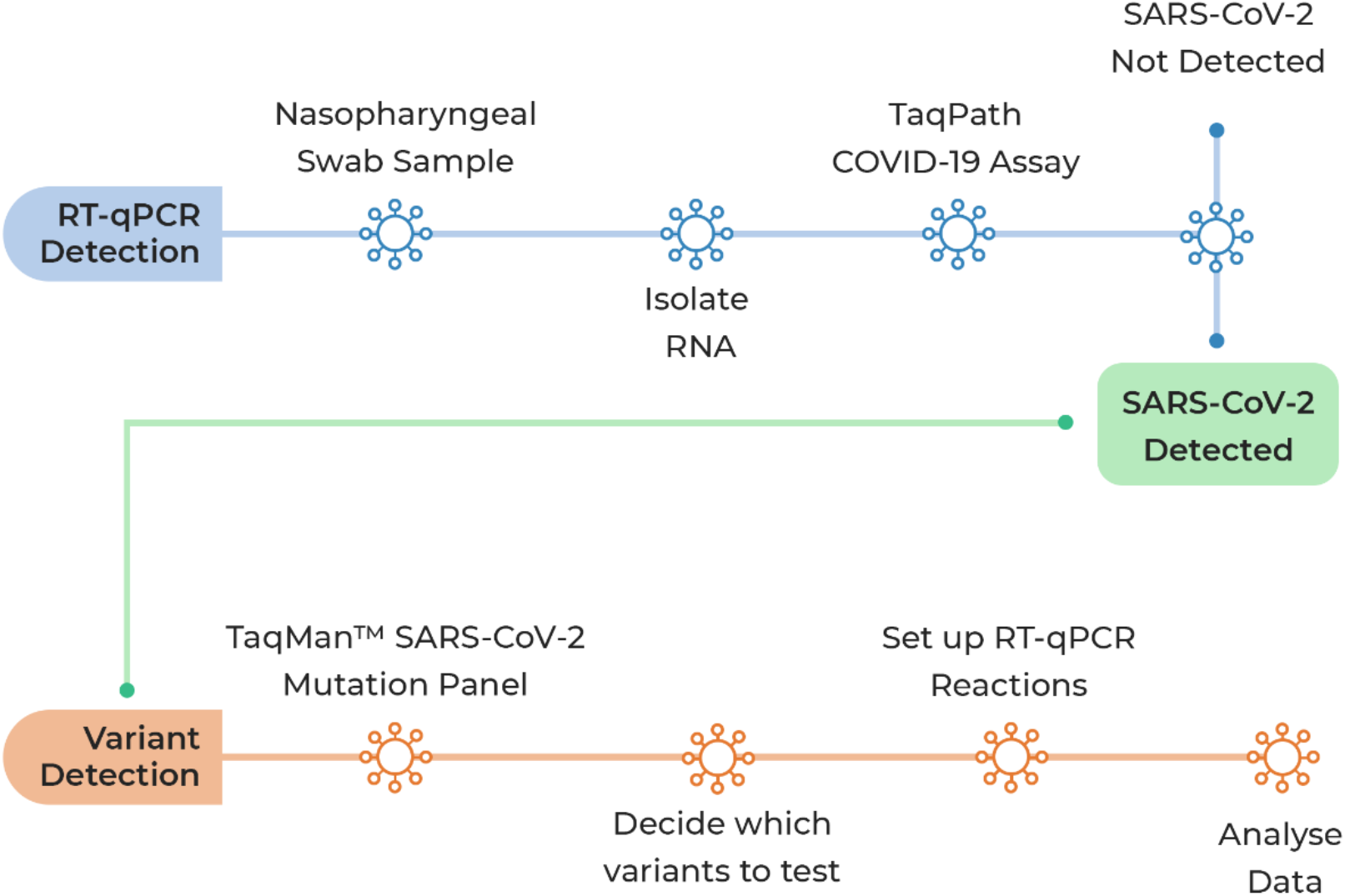
Workflow for Thermo Fisher TaqMan™ SARS-CoV-2 Mutation Panel Assay. Blue workflow: Protocol used to detect SARS-CoV-2. Orange workflow: Protocol using SNP assays to confirm mutations associated with SARS CoV-2 emerging variants.

## METHODS

### Patient samples

500 Nasopharyngeal Pillar 1 swab samples in Virus Transfer Medium (VTM) samples were sent to the University of Birmingham during April and May 2021 from Birmingham Health Trust Hospital. SARS-CoV-2 positive samples from the University of Birmingham Lateral Flow testing site were also sent and archived Pillar 2 SARS-CoV-2 samples previously identified in February 2021 at the University of Birmingham Turnkey laboratory were also utilised. The use of anonymised samples in this study was allowed under ethics gained to aid assay development (NRES Committee West Midlands - South Birmingham 2002/201 Amendment Number 4, 24 April 2013).

### RNA extraction

RNA was extracted from 200 µl of patient sample using the Thermo Fisher MagMax Viral/Pathogen II Nucleic Acid isolation kits with MagMax magnetic beads and MS-2 phage internal control, using the automated Thermo Fisher Kingfisher Flex Magnetic Particle Processor (11).

### TaqPath™ COVID-19 assay

Reactions were prepared using the Thermo Fisher TaqPath™ COVID-19 CE-IVD RT-PCR Kit protocol (**12**). RT-PCR of reactions were performed using the Applied Biosystems™ QuantStudio™ 5 Real-Time PCR Instrument. Subsequent EDT files were transferred to a computer with QuantStudio™ Design and Analysis Desktop Software v2.5.1 for analysis of exponential curves. The TaqPath™ COVID-19 assay co-amplifies three target genes: ORF1ab, N-gene and S-gene. Results were classified as positive with respect to at least 2 single-target genes (Orf1ab, N-and S-) provided the raw RT-Ct values were below 37 for single gene target signals. Bacteriophage MS2 RNA was added to each sample as an internal positive control for each sample and to monitor potential sample inhibition. The SGTF of the TaqPath™ COVID-19 CE-IVD RT-PCR Kit was considered a proxy for the presence of Δ69/70 in the S gene of SARS-CoV-2.

### TaqMan™ SARS-CoV-2 Mutation Panel Workflow

Sample inclusion for the mutation assay required RNA extracts from positive samples (Ct ≤ 30) for the TaqMan™ SARS-CoV-2 Mutation Panel workflow. From our pool of positive samples 185 were randomly selected for the mutation panel assay following the assay workflow (Figure 1). Samples containing S-gene single-target failure (SGTF) were included in the assay as long as ORF1a and N-gene Ct values were within range as there was no compromise of assay accuracy.

### Designing a Genotyping Panel for the Mutation Assay

TaqMan probes specific to SNPs found in variants known to be circulating widely within the United Kingdom around March-May 2021 were used in this study. S-gene mutations chosen were N501Y, E484K, K417N, P618R and L452R (Figure 2). Probes detect both the reference and mutation sequences of SARS-CoV-2. Reporter dye information for the TaqMan™ SARS-CoV-2 Mutation Panel is represented in the assay context sequence, which is the nucleotide sequence surrounding the mutation site in the SARS-CoV-2 reference genome (hCoV-19/Wuhan/2019; GISAID EPI_ISL_402124). The variant allele is detected by FAM™ dye and the reference allele is detected by VIC™ dye.

**Figure 2.**
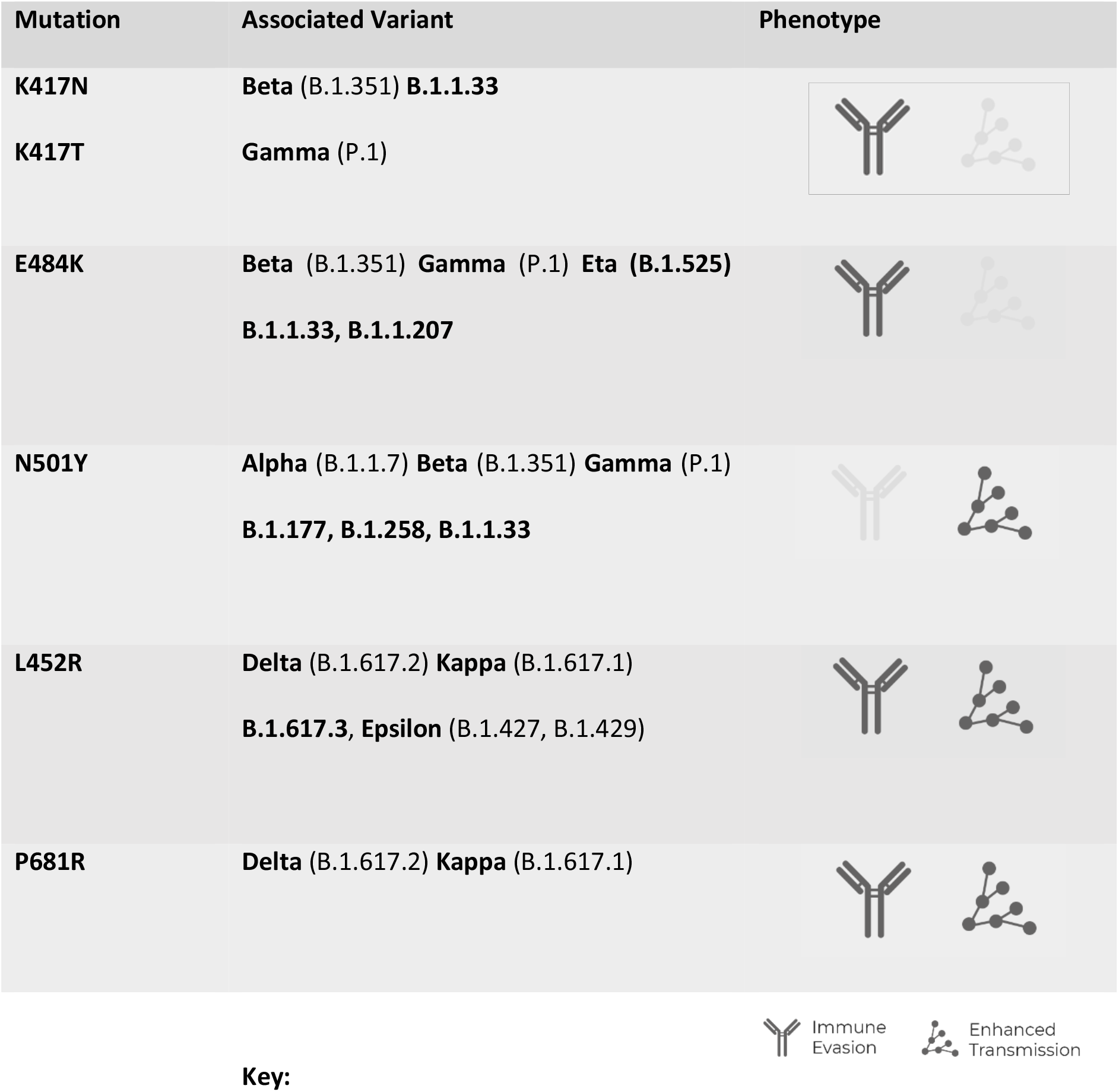
Thermo Fisher Mutation Panel Assay targets with associated SARS-CoV-2 variants and phenotype.

Presence of SGTF and N501Y was indicative of the Alpha variant, Beta variant was defined with the presence of K417N, E484K and N501Y, Gamma as E484K and N501Y without the presence of K417N, while the presence of L452R and P681R was indicative of the Delta variant.

Positive controls for both the Original SARS-CoV-2 sequence and chosen SNP mutations were used in the assay. The AcroMetrix™ coronavirus 2019 (COVID-19) RNA control (Low Positive Control), prepared using full length genomic RNA from SARS-CoV-2, was used as a positive control for SARS-CoV-2. A plasmid control containing mutation sequences for N501Y, E484K and K417N was used as a positive control for SNP mutations. However, at the time of running these experiments a plasmid control for mutations P681R and L452R was unavailable.

RT-PCR reaction mix was prepared as per the assay protocol (**12**). For samples with Ct values < 30, 5 µl of RNA was added to the reaction, for samples with Ct < 16, 2.5 µl of RNA was added to the reaction. Reactions and real-time PCR program were set up according to the mutation panel assay protocol (**13**).

### Library preparation and sequencing

Library preparation of positive SARS-CoV-2 samples (cycle threshold <30) was performed using the nCoV-2019 LoCost Sequencing Protocol version 3 (**14**), using normalised primers (New England Biolabs) for the V3 ARTIC primer scheme (ARTIC network) (**15**). Sequencing was performed on a MinION flow cell (R9.4.1) run on a GridION sequencing device (Oxford Nanopore Technologies).

The ARTIC network “nCoV-2019 novel coronavirus bioinformatics protocol” (**16**) was used to process the raw sequencing data including genome assembly and variant calling using nanopolish 0.11.3 (**17**). The genotyping was then performed using a nextflow pipeline (https://github.com/BioWilko/genotyping-pipeline). Briefly; genotypes were called using aln2type (https://github.com/connor-lab/aln2type) utilising custom variant definition files for each mutation (included in repository), and lineages were called using Pangolin (**18**).

### Data Analysis

Results were plotted as Allelic Discrimination Plots using the QuantStudio Design & Analysis v2.5 with the Genotyping Analysis Module.

## RESULTS

### Allelic discrimination plots show clear discrimination between Wild-type samples and Mutation samples using QuantStudio Design & Analysis

The Design & Analysis software genotype calling algorithm is designed for diploid organism genotype calling. The reference allele cluster should be called as homozygous allele 1, the variant allele cluster should be called homozygous_allele2 and heterozygous_allele1/allele2. Samples are called as either allele1/allele1 (ref/ref, WT), Allele2/Allele2 (mut/mut), Allele1/Allele2 (ref/mut) and no amplification (Figure 3A), thus allowing for clear and simple classification of results.

**Figure 3.**
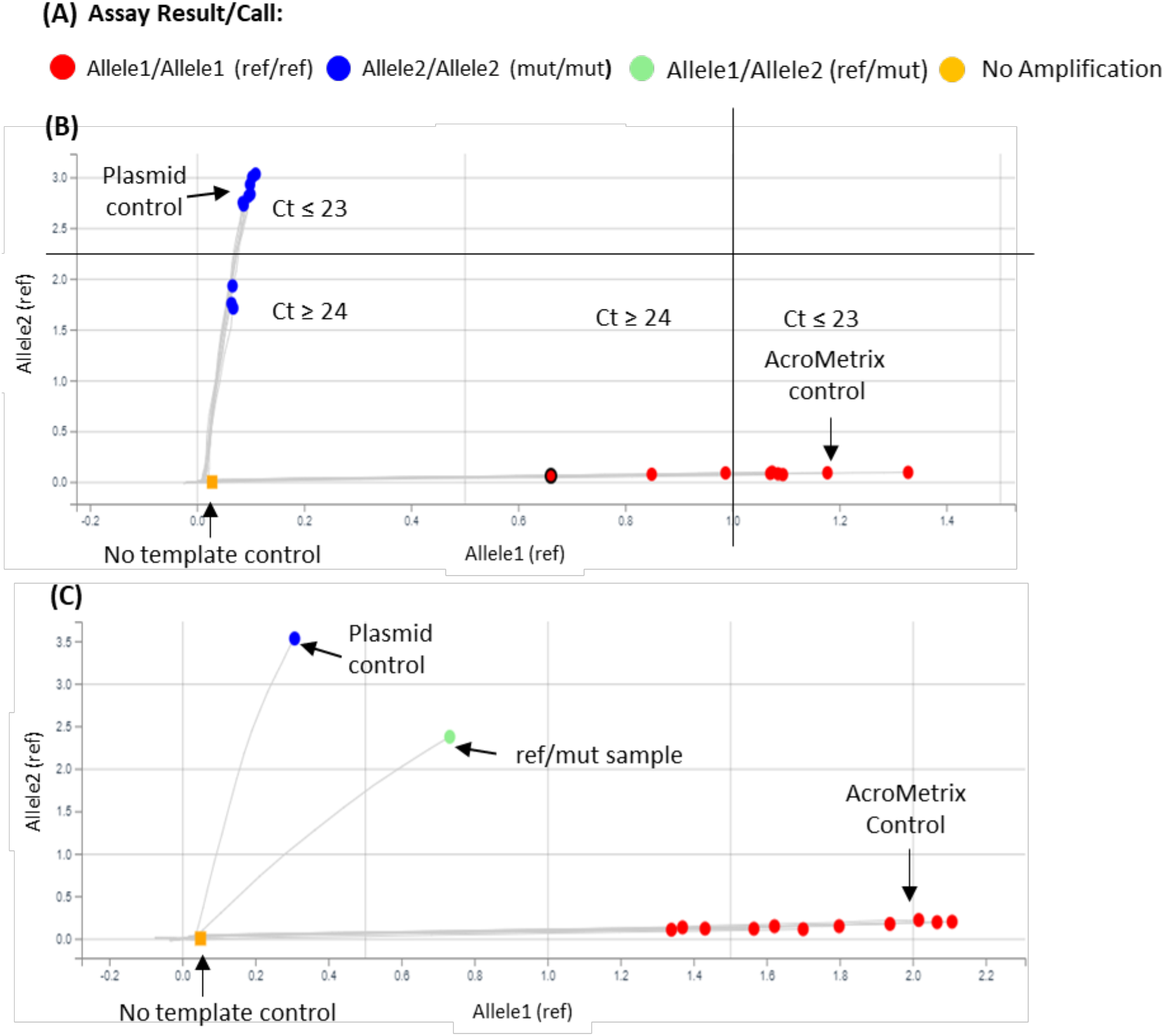
Mutation Assay Results viewed using QuantStudio Design & Analysis software with the Genotyping Analysis Module. **(A)** Assay results are ‘called’ in 4 colours according to their outcome. Red indicates allele1/allele1 (ref/ref) for WT, blue allele2/allele2 (mut/mut) for mutation present, green refers to allele1/allele2 (ref/mut) and orange showing no amplification. **(B)** Allelic discrimination plot showing clear discrimination between wild type (WT) samples (red dots along the x-axis) and the mutation samples (blue dots along the y-axis) with high and low viral loads. **(C)** Allelic discrimination plot showing an example of a ref/mut sample (green dots). Abbreviations: Ct (Cycle threshold).

Each probe is labelled with VIC dye to detect the reference (WT) sequence and FAM dye to detect the mutation sequence, which has one nucleotide difference. This allows clear discrimination on a cluster plot between WT and mutation samples, as seen with the AcroMetrix control (reference sequence) and plasmid control containing the mutation sequence, Figure 3B.

Where input samples have similar Ct values they appear as clusters on the discrimination plot, as seen in the mut/mut samples (Figure 3B) or in the case of a range of Ct values, samples are dispersed along/up the axis as seen in ref/ref samples (Figure 3B and C). We were able to detect samples with Ct’s varying from 12 to 29, in respect to ORF1ab, N-gene and S-gene. In one case we could detect mutations with Ct value 33, albeit with slightly reduced sensitivity. Samples with high viral load cluster in the upper x/y-axis and low viral load in the lower x/y-axis.

Independent mutations that are located next to one another in SARS-CoV-2 virus genome, such as P681R and P681H, can complicate genotype analysis, as probes of an assay for one mutation will fail to bind to viral sequences that contain other adjacent mutations. Mutations under the probe can appear as ref/mut and slope away from ref/ref or mut/mut samples, cluster along the X-axis, near to the NTCs, thus exhibiting weak amplification due to the probes non-specific activity, Figure 3C. Genotyping calls can manually be adjusted to ‘no amp’, or two separate assays run such as P681R and P681H to compare and facilitate accurate genotype analysis. If it is not possible to make a call, then further characterisation by genome sequencing would be necessary.

### The Mutation Panel Assay is extremely effective at identifying mutations in laboratory samples

All samples run through the mutation panel assay produced a result, with either mutation present, absent or reference/mutation, indicating another mutation within that SNP was present. Input samples had varying Ct values (Ct 12-29) with regards to each of the three single-target genes Orf1ab, N gene and S gene (Supplementary data 1). Using the Orf1ab gene as marker for Ct distribution across a Ct value group, good distribution of different Ct values was observed (Figure 4). RNA quality was not measured, and in some cases, samples had been stored at -80°C for up to 2 months and through no more than 2 freeze/thaw cycles. However, no effect on the performance of the mutation assay was observed. Furthermore, we also noted that for positive samples Ct ≤18 RNA was added at 2.5ul instead of 5ul into the Mutation Panel PCR reaction, therefore, allowing more RNA to be archived.

**Figure 4.**
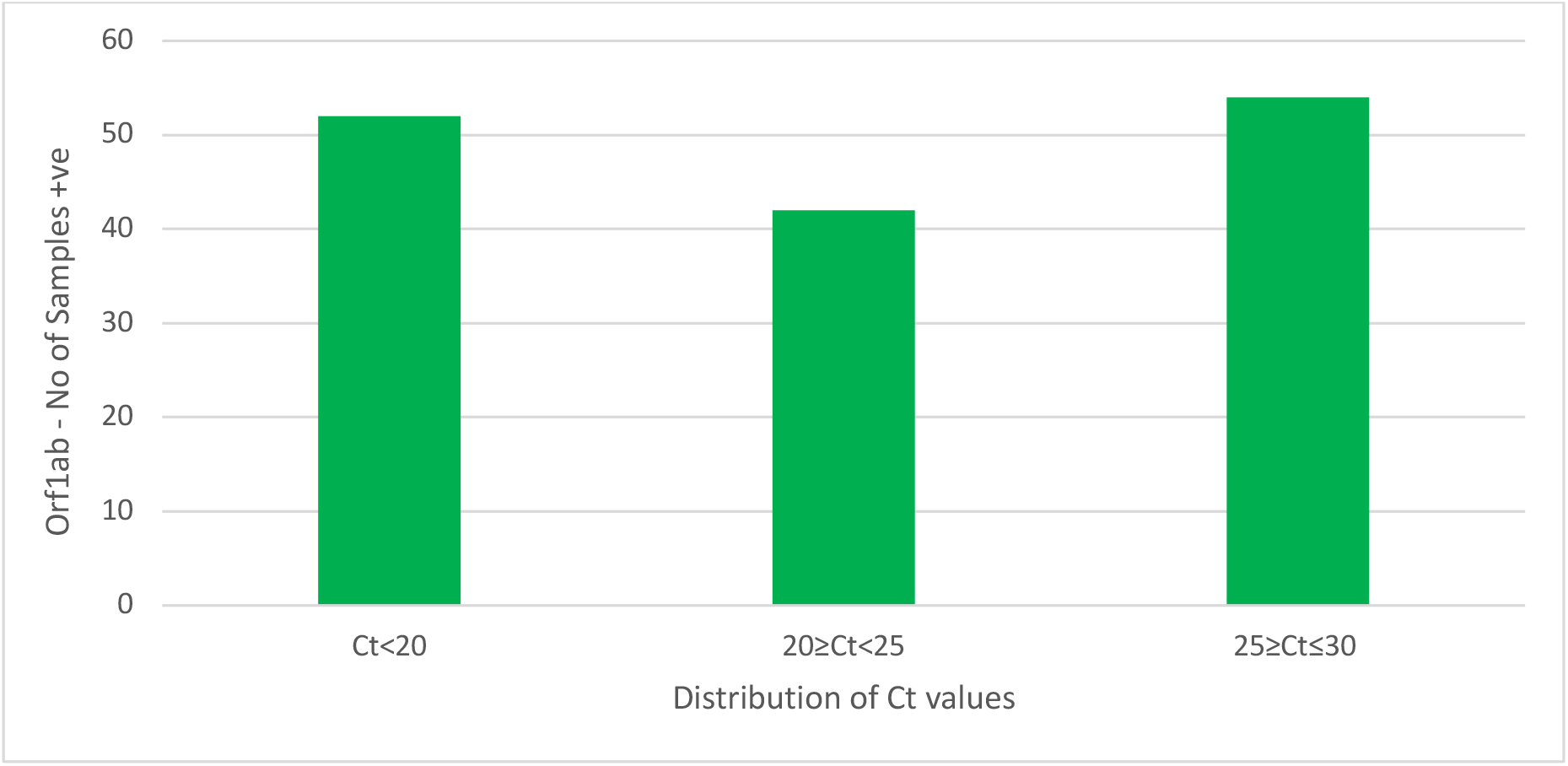
Distribution of TaqPath COVID-19 PCR Ct values for the Orf1ab gene. The Y-axis indicates the number of samples SARS-CoV-2 positive for the Orf1ab gene. The X-axis is grouped into ranges for Ct values up to and including 30. Abbreviations: Ct (Cycle threshold), Orf1ab (Open reading frame 1ab).

All samples run through this assay were sent for sequencing onsite at the University of Birmingham as part of COG-UK. This provided us with the ability to compare the mutation panel assay results with that of genome sequencing. By cross referencing the genome sequencing results for each SNP we could identify whether the mutation assay correctly calls each SNP mutation. Our data shows that for all samples, where sequencing data was available, the mutation panel is in 100% agreement (Figure 45A - Also refer to Supplementary Data 1 for complete data set for all samples run). While Nanopore sequencing may miss a SNP mutation (Figure 5B), the mutation assay can identify this. This is likely due to the RNA quality and platform used for sequencing, but highlights the importance of the genotyping assay for rapid identification and subsequent action.

**Figure 5.**
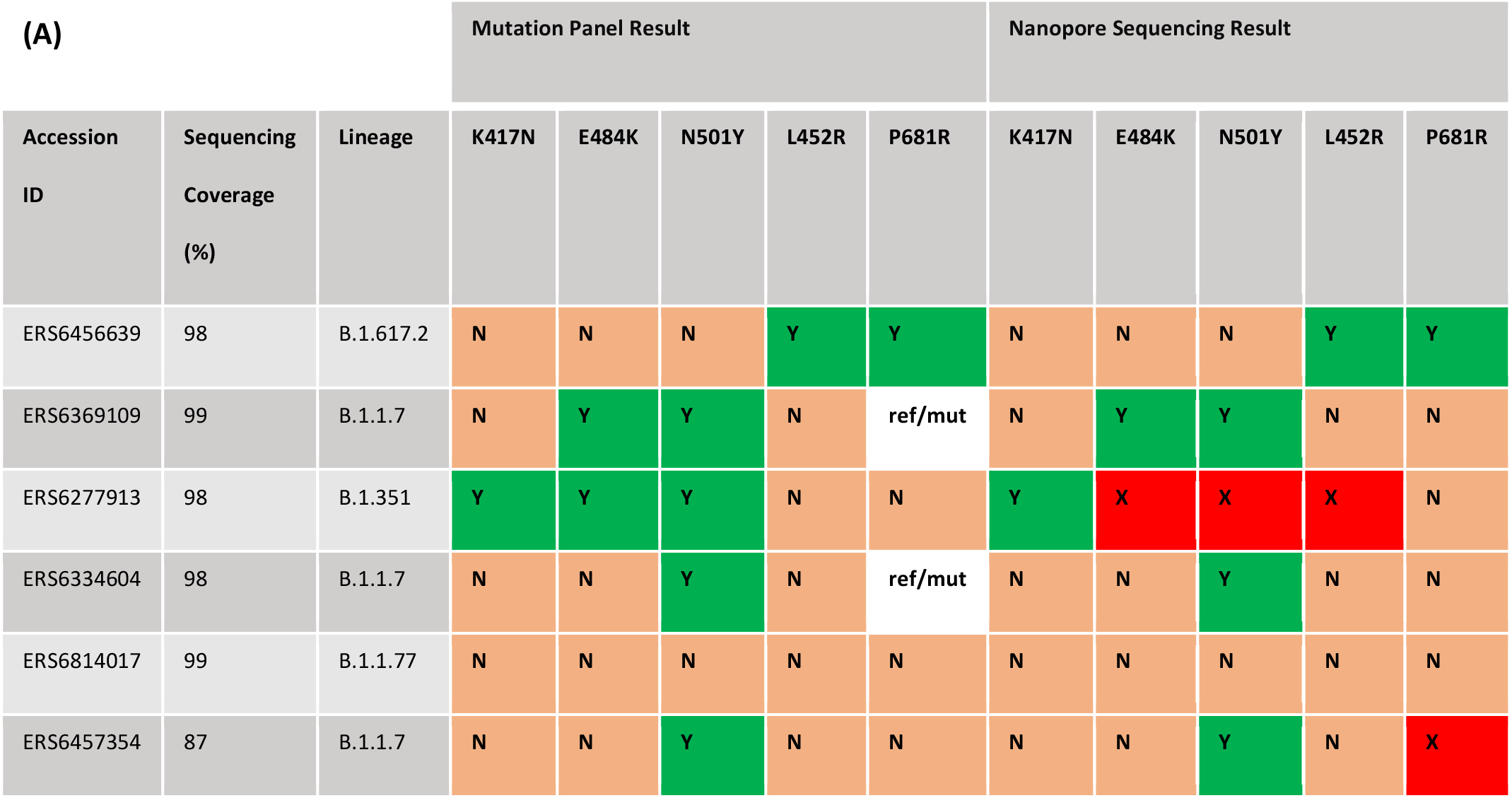
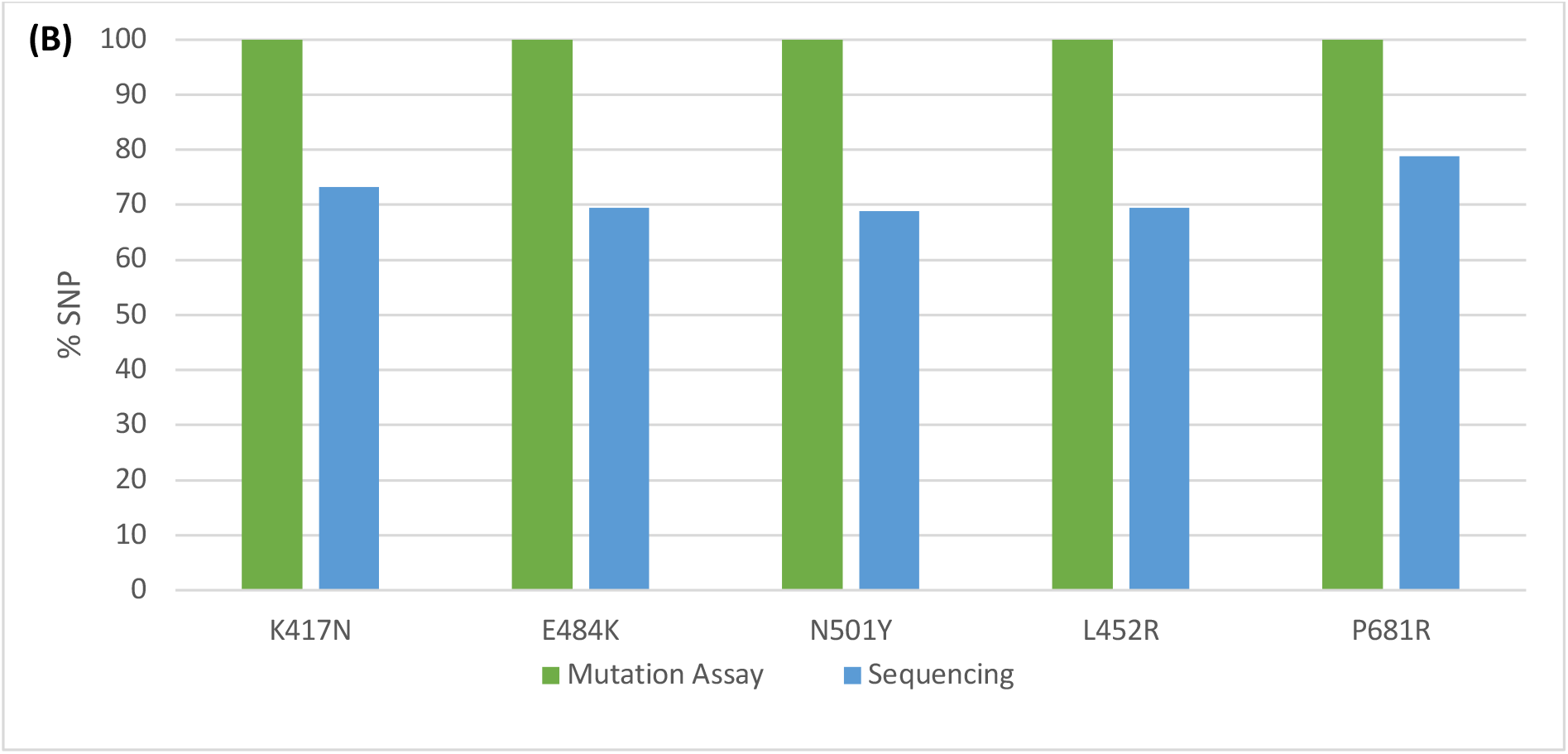
**(A)** Example of mutation assay results compared with Nanopore sequencing results from a selection of samples run through the assay. All results for all samples run are included in Supplementary Data 1. Orange Square ‘N’ = mutation not present; Green Square ‘Y’ = mutation present; White ‘ref/mut’ = mutation present on one allele only. The Red square ‘X’ indicates that there was not sufficient coverage of that SNP after sequencing. **(B)** Percentage comparison results of all mutation assay results and corresponding Nanopore sequencing data. Green Bars (Mutation Assay) = Percentage (%) SNP agreement when compared to the corresponding sequencing data. Blue Bars (Sequencing) = Percentage SNP identified when sequencing data for each sample was assigned a lineage by Pangolin.

The mutation assay cannot distinguish what the substitution is in ‘ref/mut’ results, which highlights the continued importance of sequencing and updating SNP mutations which can be added when designing an assay panel. The mutation assay cannot be used to identify variant lineages, however it can, due to the detection of specific mutations, give an indication as to which variant the sample may be and can also exclude the presence of a VOC in a sample based on the absence of key characterising mutations of significance. This is of particular importance as case numbers rise, and sequencing capacity and turnaround time may not be matched. Specifically for VOC Alpha (B.1.1.7) ref/mut further analysis of genome sequencing data revealed that this variant, although negative for P681R, was positive for P681H. Importantly, it was noted that one ref/mut was also positive for E484K and clarification from the University of Birmingham arm of the COG UK consortium confirmed that this was a small cluster of B1.1.7 variants that was being monitored and actioned in the Birmingham area.

### The Mutation Panel Assay is highly adaptable to newly emerging variants and mutations

The mutation panel designed for this assay was to identify samples containing SNPs associated with variants of SARS-CoV-2 widely circulating within the United Kingdom March 2021-May 2021, this included mutations found in the table 2. Between April and May B.1.617 variant numbers were increasing rapidly and beginning to overtake the B.1.1.7 variant within the UK population. Therefore, as experiments were being conducted in real-time the addition of P618R and L452R assays were essential for rapid identification of B.1.617 variants. Samples that had previously been run for the original assays and sent for sequencing meant that RNA availability was limited. However, freshly isolated samples from May provided the opportunity to run all assays at once.

186 samples were assayed with 182 run for N501Y, 183 for E484K and 178 for K417N, and a total of 42 samples were assayed for all 5 mutations; 68.7% of samples assayed were positive for N501Y (WT ref/ref = 31.3%); 2% of samples were positive for E484K (WT ref/ref = 97.2%); 2.8% of samples assayed were positive for K417N (WT ref/ref = 97.2%) ; 57.1% of samples were positive for L425R (WT ref/ref = 42.9%) and 61% were positive for P681R (WT ref/ref = 19%). A ref/mut ‘call’ was also noted for E484K (1 sample, 0.6%) and P681R (8 samples, 19%).

Lineage was identifiable in 76.8% (Figure 6) of samples sent for sequencing, which as mentioned previously may be due to the sequencing platform used and/or RNA quality. From our pool of samples, 65% of samples were the B.1.1.7 variant, 22% B.1.617.2, 4% B.1.177, 2% B.1.137, 1.3% B.1.351 and B.52 with B.1.177.16, B.1.1.372, B.1.1 and B.1 variants making up 2.7%. Cases of B.1.617.2 were first identified at the end of April 2021.

**Figure 6.**
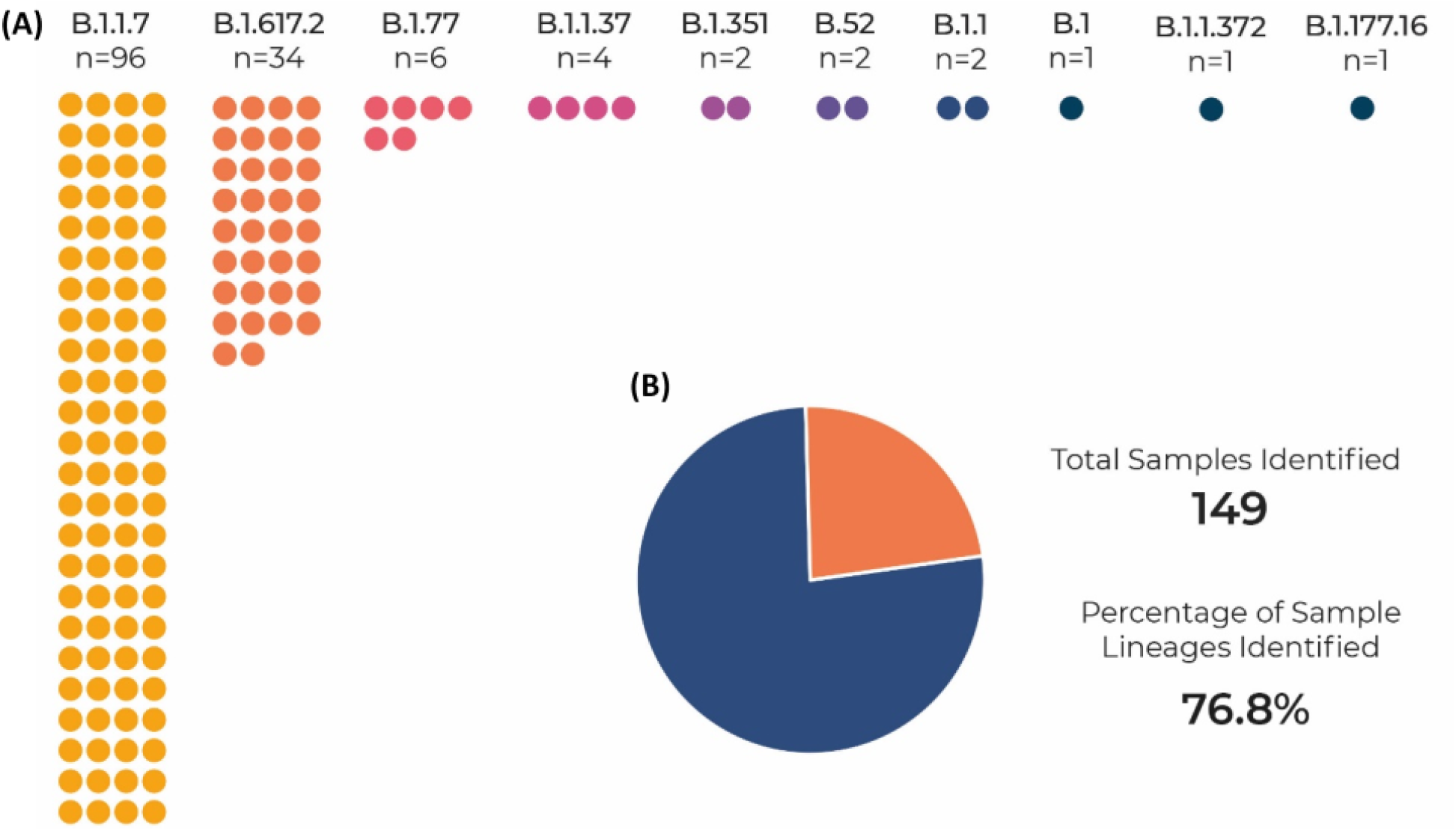
Lineage of sequenced samples identified by Pangolin. **(A)** Number of samples assigned a SARS CoV-2 lineage. **(B)** Percentage of samples assigned a lineage as compared to the total number of samples assigned a linage.

## Discussion

Genomic epidemiology is a powerful tool for tracking transmission and importation of SARS-CoV-2 as well as assessing the effectiveness of public health measures (**1, 2, 3**). Tracking transmission within the population in real-time enables laboratories to report and governments to action. Recently, Thermo Fisher have developed genotyping assays to help bridge the gap between testing capacity and sequencing capability to receive results in real-time. A previous study from our laboratory demonstrated that B.1.1.7 was associated with significantly higher viral loads (**17**) and had a genotyping assay been available at the time, this would have helped to identify the Alpha variant much quicker and identify speed and spread of infection for quicker action and containment.

In our laboratory, an opportunity arose to genotype RNA extracted in real-time from Birmingham Trust Hospital, Pillar 2 and University of Birmingham Lateral Flow, positive SARS-CoV-2 samples. Initially using three verified SNP assays from Thermo Fisher’s Applied Biosystems™ TaqMan™ SARS-CoV-2 Mutation Panel and then expanding to five to include SNPS for the delta variant, we matched genotypes for specific mutations in variants Alpha (B.1.1.7), Beta (B.1.351) and Delta (B.1.617.2).

Here we demonstrate that these mutation panels provide robust detection of VOC even if RNA is of low quality and after more than one freeze/thaw step. Importantly, a specific mutation can be identified on the same day that a nasopharyngeal swab tests positive by RT-PCR. Where sequencing data was available, the genotyping assay always matched 100% to the correct lineage.

The ref/mut function is a key to the genotyping assay as when detected this will imply an amino acid change for the specific mutation at the genomic site where the primers amplify. This is crucial in alerting to the potential rise of a new VOI (variants of interest) so it can be monitored to see its potential to become a VOC.

Our study confirms that same day rapid real-time detection of variants present in the population is very achievable - from a swab entering the lab, processing through the TaqPath COVID-19 RT-PCR and mutation assay, where results were then available in approximately 5 hours. Confirmation of mutations present and lineage identification following genomic sequencing on-site was provided in approximately 72hours, however, when on-site sequencing is not available this may increase turnaround time significantly. Rapid identification of VOCs enables test-and-trace to identify regional clusters and perform targeted testing to prevent spread of more transmissible variants. Having both PCR set-ups in our modular testing system and on-site sequencing removes the logistics, costs and time of moving samples between testing labs and sequencing labs and reporting the results.

Importantly, the genotyping panel can be updated readily as new SNP mutations are identified via genome sequencing data from COG-UK. Delta variants emerged quickly over a few weeks and during our study we were able to source very quickly, two mutation panels for the Delta variant (L452R and P681R) and on arrival were able to action on the same day.

Genome sequencing can be inconclusive for identifying key mutations found by the mutation panels but nonetheless, it is a powerful tool for identifying SARS-CoV-2 variant lineages through phylogenetic trees. However, previous studies have discussed there are limitations due to sequence drop-out when used to identify specific key mutations **(18, 19, 20, 21)**. Nanopore sequenced long-reads can be susceptible to errors where identifying specific mutations may be problematic. These allelic drop-outs potentially affect PCR-based (tile amplicon) targeted sequencing, thus resulting in incomplete genome coverage, especially at lower amounts, resulting in the loss of both 5’ and 3’ regions that fall outside primer binding positions. The presence of SNPs in the forward and / or reverse primer binding sites may lead to complete or partial lack of amplification.

For the study presented here, we demonstrated that genotyping has two major functions. 1) Genotyping is a powerful additional, more in-depth assay for identifying specific mutations and has real clinical health value allowing laboratories to report and action VOC much quicker than genome sequencing. 2) Genotyping is an excellent additional complement to the already powerful tool of genome sequencing already proven for assigning lineages via phylogenetic trees.

Our data confirms that SARS-CoV-2 genotyping is essential for real-time identification of VOC here now and tracking those that emerge for informing public health strategy.

## Data Availability

All data produced in the present work are contained in the manuscript

## Acknowledgements

Reagents and technical assistance for this study were provided free of charge by Jelena Feenstra, Tamra Hill and Manoj Ghandi at Thermo Fisher. Thermo Fisher had no influence or role in study design, data analysis, data interpretation or construction of the manuscript. The UoB Turnkey Covid diagnostic lab was created wholly from funds directly attributed by DHSC. COG-UK is supported by funding from the Medical Research Council (MRC) part of UK Research & Innovation (UKRI), the National Institute of Health Research (NIHR) [grant code: MC_PC_19027], and Genome Research Limited, operating as the Wellcome Sanger Institute.

